# Prevalence and risk factors related to *Fasciola* spp. transmission in Northern Vietnam: A cross-sectional study in multiple hosts

**DOI:** 10.1101/2024.07.10.24310192

**Authors:** Vinh Hoang Quang, Veronique Dermauw, Dung Do Trung, Binh Vu Thi Lam, Nguyen Thi Thu Hien, Nguyen Ngoc Ha, Dao Thi Ha Thanh, Brecht Devleesschauwer, Theodorus de Jong, Linda Paredis, Nathalie De Wilde, Stijn Casaert, Katja Polman, Steven Callens, Pierre Dorny, Bruno Levecke

## Abstract

**Background:** Fascioliasis, caused by *Fasciola hepatica* and *Fasciola gigantica*, is a global veterinary problem in livestock and an emerging zoonotic disease in various countries. Here we present prevalence estimates of *Fasciola* spp. in all hosts involved in the life cycle and identified risk factors associated with *Fasciola* transmission in northern Vietnam.

**Methodology/Principal findings:** We conducted a cross-sectional survey in a community in Nghe An province where fascioliasis is endemic and inhabitants commonly consume raw vegetables. Applying a simple random and cluster sampling approach, we collected 1,137 stool and 1,390 blood samples from 1,396 human participants. From 664 buffaloes and cattle, we collected 656 fecal and 534 blood samples. We also collected 330 lymnaeid snails and 233 water plant samples. Human and animal fecal samples were examined by copro-microscopy, while blood samples were screened by ELISA to detect *Fasciola* serum antibodies. We examined infections in snails using PCR and metacercariae on water plants deploying an in-house technique. Descriptive analysis and logistic regression models were applied to estimate the prevalence of and risk factors for *Fasciola* infections. While the prevalence of *Fasciola* infections was very low in humans (0% by copro-microscopy; 0.07% by ELISA), it was high in animals (52% by copro-microscopy; 54% by ELISA). In the multivariable analysis, age was the only factor associated with *Fasciola* infections in animals. *Fasciola* could not be detected in any of the sampled water plants or lymnaeid snails..

**Conclusion/Significance:** This study indicated a high prevalence of *Fasciola* infections in animals and a very low prevalence in humans in a rural community in northern Vietnam. It is recommended to implement a control program to reduce the infection rate in buffaloes and cattle. Furthermore, health education activities for livestock owners should be carried out in *Fasciola* endemic areas.

**Author Summary:** Fascioliasis is a parasitic disease, caused by the flatworms *Fasciola hepatica* and *Fasciola gigantica*, affecting the liver of livestock and humans. Although human fascioliasis is known to occur worldwide, it mainly affects the poorest communities in rural areas across subtropical and tropical countries, with recent emergence in countries such as Vietnam. We conducted a cross-sectional survey on *Fasciola* spp. infections in different hosts (e.g., humans, buffaloes, cattle, water plants, and snails) in a rural community in northern Vietnam. Our results showed a high prevalence of fascioliasis in livestock, and a very low prevalence in humans, in a rural community in Nghe An province. *Fasciola* could not be detected in the sampled lymnaeid snails and water plants. In livestock, the occurrence of *Fasciola* infection was associated with increasing age. Treatment of the community livestock population as well as specific awareness campaigns for livestock owners are suggested to reduce disease transmission.

## 1. Introduction

Fascioliasis is a parasitic disease caused by the trematode species *Fasciola hepatica* and *F. gigantica* [1]. It is one of the most important parasitic diseases of livestock globally. Today, it is estimated that 2.4 million people are infected, and that 180 million people are at risk in over 70 countries [2, 3]. The most affected countries are Bolivia, Peru, Egypt, Iran and Vietnam [3]. The World Health Organization (WHO) recommends large-scale deworming programs in endemic countries where at-risk populations are periodically provided an anthelminthic treatment [3].

Infected livestock and humans (final hosts) shed worm eggs with their stool. Once in a favorable environment (i.e., freshwater sources), miracidia (larval stage) develop in the eggs, hatch, and infect freshwater snails (intermediate hosts). In the snail tissue, the miracidia undergo several development stages (sporocysts and rediae) and various rounds of asexual multiplication. Subsequently, an exponential number of free-swimming cercariae (larval stage) are released from the snail and encyst into metacercariae (infectious stage) on water plants (the carrier). Susceptible livestock and humans acquire the infection by ingesting these contaminated raw water plants. Following infection, metacercariae excyst and the larvae migrate through the liver where they mature to hermaphrodite adults in the bile ducts and start producing eggs [4, 5].

In Vietnam, the incidence of human fascioliasis has escalated to concerning levels, prompting the WHO to classify it as a major public health concern [6, 7], and listing it as one of the countries that suffer the most from this disease [3]. Between 2014 and 2017, the annual number of human fascioliasis cases increased by a factor of four (from 2,994 to 11,706 cases; data made available by the National Institute of Malariology, Parasitology, and Entomology (NIMPE) and the Institute of Malariology, Parasitology, and Entomology Quy Nhon (IMPE-QN)). Most of these cases originate from the Red River Delta and Central Vietnam. The farming systems in rural areas in Vietnam allow livestock to freely graze, spreading *Fasciola* eggs beyond the pastures. Livestock feces is also often used or sold as fertilizer [8, 9]. These practices explain the geographical overlap in fascioliasis distribution across both humans and livestock, with the highest prevalence of *Fasciola* infection in livestock (up to 72%) being observed in the Red River Delta and Central Vietnam [7].

The aims of this study were (i) to estimate the prevalence of *Fasciola* spp. infection in the different hosts included in the lifecycle, i.e., humans, buffaloes, cattle, water plants and lymnaeid snails; (ii) to identify risk factors associated with *Fasciola* infections in a rural community in Northern Vietnam. The results from this study are important for updating information on fascioliasis epidemiology and developing effective control strategies in highly endemic areas.

## 2. Methods

### 2.1. Ethics statement

This study was reviewed and approved by the institutional review boards of both NIMPE in Vietnam (No. 02-2022/HDDD), and the Ghent University Hospital/Faculty of Medicine and Health Sciences, Ghent University in Belgium (No BC-08915). Before the start of the fieldwork, we informed the provincial and district health officers, and the Dong Thanh Commune People’s Committee (including the commune medical doctors and veterinarians) about the study aims and procedures, which subsequently also approved the study. Detailed information on the study objectives and procedures was provided to each participant on the first visit, and a signed informed consent from each participant aged 10 years and older was obtained before enrolment. Children between 5 and 18 years old were asked to verbal assent, and their parents were invited to provide a written informed consent. Any individual that tested positive for fascioliasis based on any of the applied laboratory tests was informed and advised to consult a medical doctor at the Provincial General Hospital or the NIMPE hospital for further examination. For the animal study, we obtained permission from the owners to collect fecal and serum samples of their livestock.

### 2.2. Study site

This study was conducted in Dong Thanh commune of the Yen Thanh district in the Nghe An province in Northern Vietnam. The selection of this study site was based on (i) historical data on the presence of fascioliasis, (ii) cultural behaviors that favor disease transmission (i.e. consumption of raw vegetables), (iii) the presence of both buffaloes and cattle that have access to crop fields (e.g., rice and water plants) and (iv) the accessibility of the study site. Nghe An province consists of 21 districts of which Yen Thanh has the second-highest number of human fascioliasis cases (between 2012 and 2020, 398 cases were reported by the Center for Disease Control of Nghe An province).

Yen Thanh district consists of 39 communes of which Dong Thanh commune had the highest number of positive cases of fascioliasis (between 2012 and 2020, 39 cases were reported by the Center for Disease Control of Nghe An province). This commune has 8,490 registered inhabitants across 2,036 households. The main occupation of residents in this commune is farming. In total, there were 873 buffaloes and cattle, which were raised by 452 households. In this commune, there were 370 hectares of rice fields and a canal system to irrigate the fields.

### 2.3. Study design

Between April and May 2022, we conducted a cross-sectional study both, (i) to estimate the prevalence of *Fasciola* spp. in different hosts, including humans, buffaloes, cattle, and lymnaeid snails, and on water plants, and (ii) to identify putative risk factors for *Fasciola* spp. infections in the Dong Thanh commune. In the following sections, we will discuss each of the study populations in more detail.

#### 2.3.1. Human population

A list of all households was provided by the Dong Thanh Commune People’s Committee. Based on this list 623 households were randomly selected using R software [10]. During the first house visit, eligible household members were invited to participate, and subsequently asked to provide a stool and blood sample and to complete a structured questionnaire. People were eligible when they met the following inclusion criteria (i) being a Dong Thanh commune resident or having been in the commune for at least six months at the first visit, and (ii) being at least five years old. Whenever possible, three eligible household members were invited to participate. In case more than three people were present in the household, we randomly selected three participants. Only the selected individuals that signed an informed consent form, and in case of children, provided verbal assent, participated in the study. Each participant was interviewed using an individual questionnaire to obtain demographic characteristics, and knowledge, awareness, attitudes, and practices of fascioliasis (see **Info S1** for a template of the questionnaire). One household member was also interviewed to obtain general information on water and sanitation practices, livestock and crop management, and culinary practices (see **Info S2** for a template of the questionnaire). Subsequently, venous blood samples were collected by health staff. Each participant was given a pre-labeled plastic container, and they were asked to provide a stool sample by the next day (visit 2). These stool samples were collected by health workers or during the third visit when water plant samples were collected (see section 2.3.4 Water plants). Participants who did not provide stool samples during these two consecutive visits were asked to bring the sample to the commune health station.

#### 2.3.2. Buffalo and cattle population

A list of all households that raised water buffaloes (*Bubalus bubalis*) and cattle was provided by the local veterinarian. The research team visited each of the households on this list. On arrival, the owner was informed about the study’s aims and procedures. Only animals for which the owner assented were included in the study. Both a venous blood sample and a rectal feces sample were collected from each animal. In addition, general information about the animals was collected, including number of animals owned by the household, age, sex, and animal species (buffaloes *vs.* cattle).

#### 2.3.3. Snail population

In total, five different snail habitats (vegetable field, irrigation canal, rice field, shallow pond, and lake) were identified by the village head. At each habitat, two team members searched for snails for 15 minutes. All snails were collected while recording the habitat of the snails. Upon arrival at the commune health station, the collected lymnaeid snails were initially identified as by Doanh et al. [11], and stored in containers with 70% ethanol on a species and habitat basis. Finally, these containers were sealed by parafilm and transferred to the Institute of Tropical Medicine, Antwerp (Belgium) for molecular analysis.

#### 2.3.4. Water plants

At the first visit, households were instructed to collect water plants in the three following days (see **Info S3** for a template of the instructions). For this, we provided them with a plastic bag with a unique household ID number and asked them to collect about 300 grams of freshwater plants that they would consume either raw or undercooked. The samples were collected by health workers of villages and delivered to the research team at the commune health station. Next, these samples were kept at 4 °C before being transferred to NIMPE for further analysis within three days.

### 2.4. Sample size calculation

For the sample size calculation of humans, we ran simulations in the statistical software R (2020) [10], envisaging a probability of 95% to attain the apparent prevalence and a maximum precision of 5% for a prevalence estimate for each of the two study populations [12]. To account for clustering of the human study populations, we considered a design effect of 3 [13]. Based on the reported prevalence (humans: 7.8%) [14], a total of 1,381 humans were aimed to be sampled. For buffaloes/cattle we aimed to sample all animals present in the commune. For the water plants, we asked each selected households in the human study to provide freshwater plant samples. Finally, all lymnaeid snails were searched across all habitats for a duration of 15 minutes.

### 2.5. Laboratory analysis

#### 2.5.1. Stool samples

Both human and animal stool samples were processed within 24 hours of collection at the commune health station. All stool samples were examined microscopically for *Fasciola* spp. eggs by means of the commercially available Fluke Catcher (Provinos). The method was shown to be the most sensitive and precise technique for *Fasciola* diagnosis in human stool, as compared to other techniques such as Kato-Katz and Mini-FLOTAC [15]. For the animal samples, eight grams of stool were processed for each animal separately, while for human samples two grams of four individual stool samples were pooled. In case *Fasciola* eggs were observed in these pools, the individual stool samples that formed the pool were individually processed. This cascaded pooled test strategy for human samples was preferred, as the prevalence was expected to be low. To obtain the fecal egg counts (FECs; expressed as eggs per gram stool (EPG)), the raw egg counts were divided by eight and two for animal and human samples, respectively. The presence of any helminth egg other than *Fasciola* (e.g., *Paramphistomum* in animals) was recorded. **Info S4** provides a detailed standard operating procedure (SOP) for both animal and human samples.

#### 2.5.2. Serum samples

Collected blood sample tubes were placed in a fridge (4 °C). The next day, the clotted blood was separated by centrifugation at 3,000 rpm for 15 min, and the serum was dispensed into pre-labeled vials, and stored at -20 °C at the commune health station. Once the fieldwork was completed, all the serum samples were transferred to NIMPE in a cool box. At NIMPE, aliquots were frozen at -86 °C until further analysis. The serum samples were examined for exposure to fascioliasis by means of a commercial antibody (Ab)-ELISA (BioX K211, BioX, Rochefort, Belgium). The optical density ratio (ODr in %) was calculated by multiplying the optical density (OD) of the sample by 100 and dividing it by the OD of the positive control. A sample was considered positive when the OD ≥ 15%, while values less than 10% were considered negative. In all other scenarios, the test result was considered inconclusive. This test was developed to detect fascioliasis in cattle and sheep. For the human samples, the test was modified by using an anti-human IgG and by standardization of the test result with positive human sera [16]. **Info S5** provides a detailed SOP of this Ab-ELISA for both animal and human samples.

#### 2.5.3. Snails

For the DNA extraction, individual snails were crushed in a 100 μL 5% CHELEX solution and we added 0.015 μL PhHV-1 solution (Phocid Herpes Virus subunit 1, kindly provided by the European Virus Archive GLOBAL) to control for inhibition of the PCR reaction. Then, the samples were incubated one hour at 56 °C and for 30 min at 95 °C. Afterwards the mixture was centrifuged for 7 min at 13,000 *g*. The supernatant containing the DNA was subjected to a multiplex PCR amplifying DNA of both an ITS1 segment of *Fasciola* spp. (716 bp) and the gB gene of PhHV-1 (89 bp). The PCR reactions were performed in a 25 µL volume containing 1x Qiagen® Multiplex PCR mix, 0.25 µL template DNA, 0.25 µM Fasc-ITS1 F (5’ TCT ACT CTT ACA CAA GCG ATA CAC 3’) and Fasc-ITS1 R primers (5’ GGC TTT CTG CCA AGA CAA G 3’) [17] and 0.375 µM PhHV-1 F (5’ GGG CGA ATC ACA GAT TGA ATC 3’) and PhHV-1 R primers (5’ GCG GTT CCA AAC GTA CCA A 3’) [18]. The reaction mixture was run in a Biometra Thermal Cycler for 5 min at 95 °C, 40 cycles of 30 sec at 95°C, 30 sec at 55 °C and 30 sec at 72 °C and ending with a final elongation at 72°C for 5 min. The PCR products were run on a 2% agarose gel for 30 min at 100 V and visualized by EtBr staining and UV light illumination. For samples that did not present the 89 bp inhibition control, the PCR reaction was repeated on dilutions of the DNA until a fifth of the original concentration samples that still did not present the control sequence were disregarded.

#### 2.5.4. Water plants

Water plant samples were immediately processed upon arrival at NIMPE. As samples were collected over three consecutive days, representing different water plants, we weighed the sample and divided it over four piles. Then, 25 gram of each pile was transferred into a plastic ziplock bag of 5 L. Subsequently, the samples were processed by applying an in-house validated laboratory technique [19]. In brief, 1 L of tap water was added to the plastic bag, which was closed and put into a 10 L ziplock plastic bag to avoid leakage. The bag was put in a concrete mixer for 10 min. Subsequently, the sample was sieved over a tower of three metal sieves (30 cm x 3 cm in diameter) each with a different mesh size (from top to bottom: 500 µm, 350/355 µm, 100 µm). To enhance recovery of the metacercariae, plant material was sieved in two steps. To be more explicit, half of the sample was transferred on the sieves, while thoroughly washing the material. Then this half was discarded, and the second half was sieved and washed. In addition, the ziplock bag was thoroughly rinsed, and the content was transferred over the tower of sieves. To further reduce the volume, the material on the 100 µm sieve was recovered and sieved over a pluriStrainer sieve (diameter: 3 cm, mesh size: 150 µm). Then, the material on the pluriStrainer sieve was collected in a small recipient by inverting it and washing/rinsing it with tap water using a 1 mL pipet. Finally, the entire content was screened for the presence of metacercariae using a stereomicroscope. To this end, parts of the content was transferred into a petri dish and screened, after which the petri dish was washed and filled again. When metacercariae were detected, they were transferred to a 1.5 mL tube containing 70% ethanol. These preserved samples were stored at room temperature to confirm the presence of *Fasciola* metacercariae based on a molecular analysis [19].

### 2.6. Statistical data analysis

The statistical data analysis was conducted in three consecutive steps. First, we conducted a descriptive analysis of the data, separately summarizing the demographic characteristics of different study populations based on host species, age, sex (all), education, and occupation (in humans only). The apparent prevalence was estimated for each diagnostic test separately by dividing the number of positive cases by the number of sampled cases. The “binom” package in R was utilized to estimate the prevalence of *Fasciola* infection and its associated Wilson or exact (when cell counts were less then five) 95% confidence interval (95% CI). The intensity of infections based on copro-microscopy was classified into light (0 EPG < FEC ≤10 EPG), moderate (11 EPG < FEC < 25 EPG), and heavy (≥25 EPG) [20]. Generalized mixed models were built to identify putative risk factors for *Fasciola* infections in livestock, with the test result on copro-microscopy (absence *vs.* presence of eggs) as outcome variable and host species (buffaloes *vs.* cattle), age (in months), sex (bull *vs.* cow), and sources of feed and drinking water as predictive variables, while accounting for household as a random effect. The sources of feed and drinking water were captured through questionnaires (see **Info S2**). For the source of feed, seven variables were included: cut and carry, growing in or near water bodies (yes *vs.* no); cut and carry, grown elsewhere (yes *vs.* no); tethering (yes *vs.* no); factory product (yes *vs.* no); free roaming/grazing, in or near waterbodies (yes *vs.* no); free roaming/grazing elsewhere (yes *vs.* no); other (yes *vs.* no). For the source of drinking water, seven variables were included (piped water into dwelling (yes *vs.* no); tubewell/borehole (yes *vs.* no); protected dug well (yes *vs.* no); rainwater collection (yes *vs.* no); surface water (yes *vs.* no); other (yes *vs.* no); do not know (yes *vs.* no). The model buildup was conducted in three consecutive steps. First, we performed a univariate analysis, where variables resulting in a *p*-value <0.20 were considered in the next step. Second, the main effects of these variables were explored. Variables not significantly (*p*-value >0.05) explaining the presence of eggs in stool (‘ANOVA’ function with type III test), were omitted from the model. In a final step, two-way interactions between the remaining variables were explored. For this, we compared the main effect model with a two-way interaction model (‘ANOVA’ function, Chi^2^ test). Due to a poor agreement between copro-microscopy and Ab-ELISA for buffaloes (suggesting that the Ab-ELISA kit is not appropriate for this host species), we restricted the analysis to copro-microscopy only. Putative risk factors for human *Fasciola* spp. were not explored as the prevalence was too low (copro-microscopy: 0% and Ab-ELISA: <1%). The data were analyzed using R software version 4.3.3 for Windows [21].

## 3. Results

### 3.1. Prevalence of *Fasciola* infection in humans

A total of 1,396 human individuals participated in the study, of which the majority were females (55.30%). A quarter of the participants (25.50%) were below the age of 18, and half were between 18 and 59 years of age (18 to 39 years: 21.85%; 40 to 59 years: 35.46%). Another 17.12% were at least 60 years old. In a negligible percentage (0.07%), the age was not recorded. The educational level varied across the participants. Half of the participants finished high school (53.94%), while 21.70% had either primary school or no education. Another 17.34% completed secondary school. A small fraction had no formal education (1.79%) or a vocational or bachelor’s degree (5.23%). Most of the participants were farmers (59.31%), underscoring the agricultural character of the region. Other occupational categories included government service or civil servants (3.44%), workers (4.80%), school children (26.29%), and individuals with various other occupations (6.09%). For a marginal percentage (0.07%) the occupation was not recorded.

Out of the 1,396 individuals from 623 households that participated in this study, 1,131 individuals across 520 households provided both blood and stool. A stool or a blood sample alone were provided by 6 participants (6 households) and 259 participants (103 households), respectively. *Fasciola* prevalence was found to be extremely low. Testing of 1,390 human serum samples with Ab-ELISA revealed the presence of antibodies in just one sample, yielding an apparent prevalence of 0.07% (95% CI: 0.00 – 0.40%). Similarly, copro-microscopy of 1,137 human stool samples detected no *Fasciola* egg (95%CI: 0.00 – 0.32%). Given this low prevalence of *Fasciola* infections, we did not pursue risk factor analysis.

### 3.2. Prevalence, intensity, and risk factors of *Fasciola* infection in buffaloes and cattle

#### 3.2.1. Prevalence

In total 664 animals from 415 households were included in this study, of which 446 (67.17%) were cattle and 218 (32.83%) were buffaloes. Majority of the animals were cows (77.41% *vs*. 22.59%). The age of the animals ranged between 6 and 200 months. In total 534 (80.42%) serum samples and 656 (98.80%) stool samples were collected. Testing of 534 livestock serum samples with Ab-ELISA revealed *Fasciola* antibodies in 289 samples, resulting in a prevalence of 54.12% (95% CI: 49.88 – 58.30%). Similarly, based on copro-microscopy *Fasciola* eggs were detected in 338 out of the 656 livestock stool samples, resulting in a prevalence of 51.52% (95% CI: 47.70–55.33%). The prevalence of *Fasciola* spp. according to copro-microscopy and Ab-ELISA in the different livestock species is presented in **Table 1**. The *Fasciola* prevalence by copro-microscopy was similar in buffaloes (53.74%) and cattle (50.45%). In contrast, the seroprevalence was markedly lower in buffaloes (8.67%) when compared to cattle (75.90%). In cattle, the prevalence based on copro-microscopy was higher in cows than in bulls (56.09% *vs.* 28.09%), while there was no difference in prevalence among sexes in buffaloes. In both livestock species prevalence based on copro-microscopy increased with age.

**Table 1.** The prevalence of *Fasciola* spp. in buffaloes and cattle in the Dong Thanh commune based on copro-microscopy and Ab-ELISA.

|  | Buffaloes |  |  |  | Cattle |  |  |  |
| --- | --- | --- | --- | --- | --- | --- | --- | --- |
|  | Copro-microscopy |  | Ab-ELISA |  | Copro-microscopy |  | Ab ELISA |  |
|  | Prevalence (%) (n/N) | 95% CI | Prevalence (%) (n/N) | 95% CI | Prevalence (%) (n/N) | 95% CI | Prevalence (%) (n/N) | 95% CI |
| **Sex** |  |  |  |  |  |  |  |  |
| Cow | 52.94<br>(81/153) | 45.06-60.68 | 7.63<br>(9/118) | 4.06-13.86 | 56.09<br>(198/353) | 50.88-61.17 | 79.5<br>(221/278) | 74.36-83.82 |
| Bull | 55.74<br>(34/61) | 43.30-67.49 | 10.91<br>(6/55) | 5.10-21.83 | 28.09<br>(25/89) | 19.81-38.18 | 63.86<br>(53/83) | 53.12-73.37 |
| **Age (months)** |  |  |  |  |  |  |  |  |
| ≤12 | 31.58<br>(12/38) | 19.08-47.46 | 16.67<br>(6/36) | 7.87-31.89 | 19.4<br>(13/67) | 11.71-30.42 | 53.85<br>(35/65) | 41.85-65.41 |
| 13 to 36 | 58.14<br>(50/86) | 47.58-68.00 | 9.21<br>(7/76) | 4.53-17.81 | 45.77<br>(92/201) | 39.03-52.67 | 76.07<br>(124/163) | 68.97-81.97 |
| 37 to 60 | 53.57<br>(15/28) | 35.81-70.47 | 9.09<br>(2/22) | 1.12-29.16 | 68.82<br>(64/93) | 58.81-77.33 | 85.51<br>(59/69) | 75.34-91.93 |
| >60 | 61.29<br>(38/62) | 48.85-12.42 | 0<br>(0/39) | 0.00-9.03 | 66.67<br>(54/81) | 55.85-75.97 | 87.5<br>(56/64) | 77.23-93.53 |
| **Total** | **53.74**<br>**(115/214)** | **47.05-60.29** | **8.67**<br>**(15/173)** | **5.32-13.81** | **50.45**<br>**(223/442)** | **45.81-55.09** | **75.9**<br>**(274/361)** | **71.23-80.02** |

**Table 2** compares the results of copro-microscopy and Ab-ELISA for the detection of *Fasciola* infections. In buffaloes, there was an agreement in test results (positive/positive or negative/negative/suspected) in 67 out of the 169 animals (39.65%). In the remaining 102 samples, the majority (43.20%) of the animals tested positive on copro-microscopy but negative on Ab-ELISA. In cattle, there was a better agreement in test results (64.43% *vs.* 39.65%). In the remaining animals, 1.68% tested positive on copro-microscopy and negative on Ab-ELISA, while 31.09% tested positive on Ab-ELISA but negative on copro-microscopy.

**Table 2.** Comparison in test results across copro-microscopy and Ab-ELISA for the detection of *Fasciola* infections in buffaloes and cattle.

| Ab-ELISA | Copro-microscopy | Buffaloes |  | Cattle |  |
| --- | --- | --- | --- | --- | --- |
|  |  | n | % | n | % |
| Negative | Negative | 58 | 34.32 | 69 | 19.33 |
| Negative | Positive | 73 | 43.20 | 6 | 1.68 |
| Suspected | Negative | 12 | 7.10 | 7 | 1.96 |
| Suspected | Positive | 11 | 6.51 | 3 | 0.84 |
| Positive | Negative | 6 | 3.55 | 111 | 31.09 |
| Positive | Positive | 9 | 5.33 | 161 | 45.10 |
| **Total** |  | **169** | **100%** | **357** | **100%** |

#### 3.2.2. Infection intensity

Overall, the majority (59.47%) of animals had low intensity infections, followed by moderate infections (18.34%). Heavy intensity infections were observed in 22.19% of the animals. Across the buffaloes and cattle, the proportion of low infections was 60.00% and 59.19%, respectively. **Fig 2** further illustrates the infection intensities across age groups (**Fig 2A**) and sex (**Fig 2B**) for each of these animal species separately. Generally, the figure indicates some important differences between animal species. While in cattle the proportion of moderate-to-heavy intensity infections increase with age and are highest in bulls, in buffaloes there was a peak in the moderate-to-heavy intensity infections in animals between the age of 37 and 60 months, and no differences across sexes were observed.

**Fig 1.**
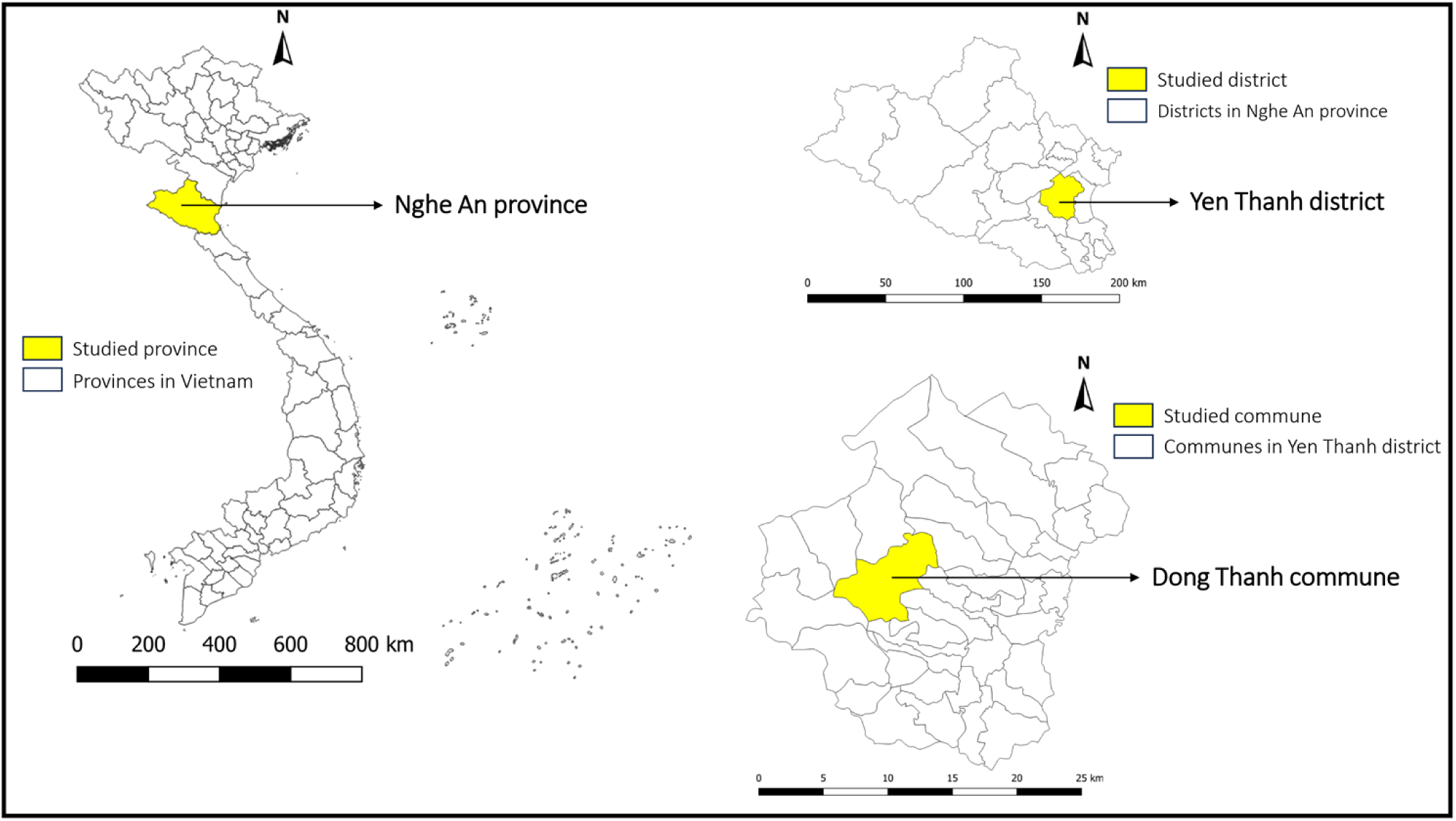
Map of Vietnam showing the locations of Nghe An province, Yen Thanh district, and Dong Thanh commune. The map was created using QGIS Desktop version 3.36.2 software

**Fig 2.**
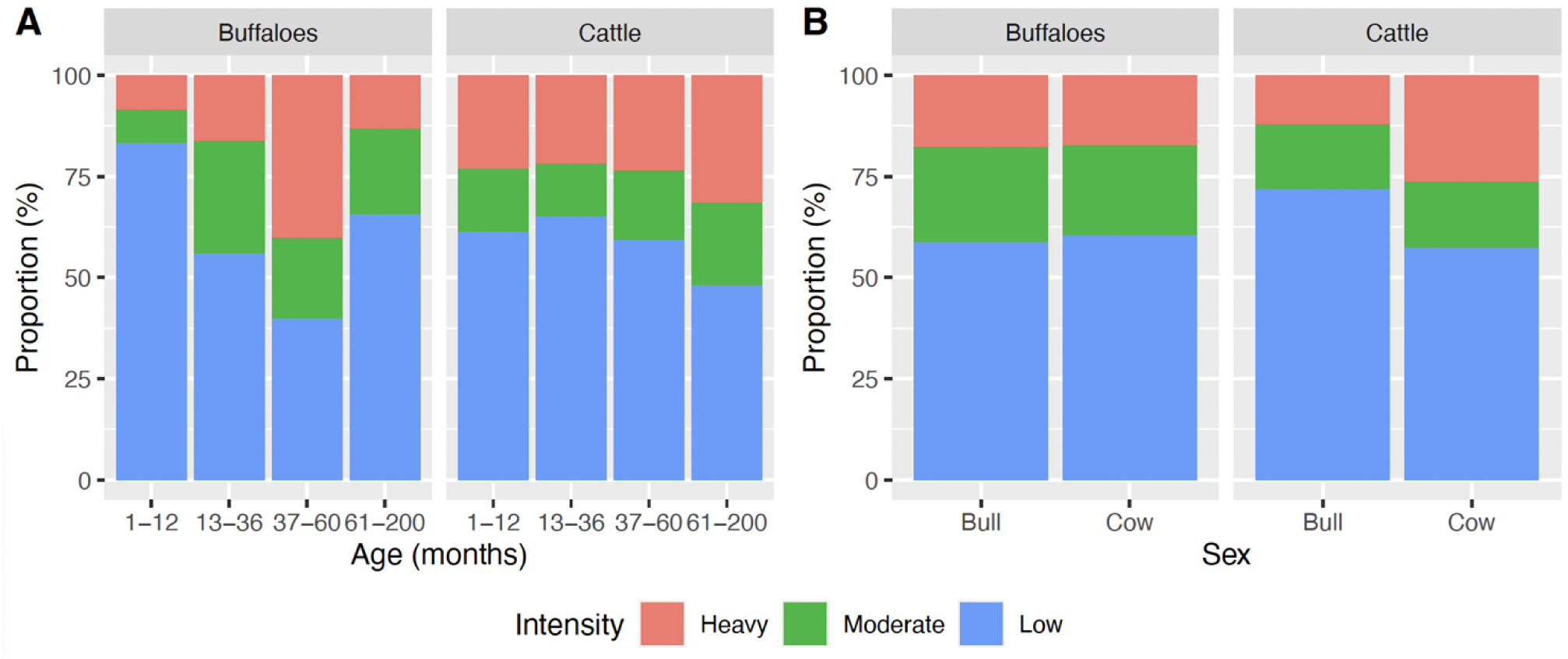
The intensity of *Fasciola* infections across age groups and sex according to copro-microscopy for buffaloes and cattle. These bar plots represent the infection intensity measured by copro-microscopy across age groups **(Fig2A)** and sex **(Fig 2B)** for buffaloes and cattle.

#### 3.2.3. Risk factors for *Fasciola* infections in buffaloes and cattle

The univariate analysis revealed that sex (*p* = 0.020), age (in months; *p* = 0.006) and feeding by cut and carry, grown in or near water bodies (*p* = 0.078) impacted the *Fasciola* infections. For all other variables the *p*-value exceeded 0.20. In the multivariable analysis, only age resulted in a *p*-value <0.05, highlighting that the odds of a *Fasciola* infection was 1.00 (95% confidence interval: 1.00; 1.02; *p* = 0.006) times higher when an animal was 1 month older. Putting this in a different perspective, the odds of infections in animals 60 months of age (75^th^ quantile) is 1.18 higher than that of animals that are 18 months of age (25^th^ quantile).

### 3.3. Prevalence of *Fasciola* infection in snails

In total, 330 lymnaeid snails were collected from 38 different locations. These locations encompassed a variety of habitats, including nine vegetable fields, nine irrigation canals, nine rice fields, eight shallow ponds, and three lakes, spread across nine villages. Molecular analysis of the collected snails was performed to detect the presence of *Fasciola* infections. Three snails were disregarded because of inhibition of the PCR reaction, noticed by the absence of amplification of the inhibition control PhHV-1. On the remaining 327 lymnaeid snails, the analysis did not reveal any *Fasciola* infections, resulting in a prevalence of 0.00% (95% CI: 0.00–1.12%).

### 3.4. Prevalence of *Fasciola* metacercariae on plants

A total of 623 households were visited, out of which 287 provided raw vegetable samples they commonly consume within three days after the household visit. Among these samples, 54 were identified as non-water plants, and were therefore discarded from any further analysis. The remaining 233 water plant samples were meticulously examined, yet no *Fasciola* metacercariae were detected in any of the samples, indicating a prevalence of 0.00% (95% CI: 0.00–1.57%).

## 4. Discussion

Human fascioliasis has escalated to concerning levels in Vietnam, prompting the WHO to classify it as a major public health concern [3]. As the disease transmission involves multiple hosts, it is of utmost importance to gain insights in the disease occurrence and associated risk factors, with the goal to design control strategies. In the present study, we estimated the prevalence of *Fasciola* spp. infection in all hosts included in the lifecycle and identified putative risk factors in a rural community in Northern Vietnam where (i) historical data show the highest number of cases of human fascioliasis, (ii) cultural behaviors are practiced that favor disease transmission, and where (iii) both cattle and buffaloes have access to crop fields. Our results showed a high prevalence of *Fasciola* infection in livestock, but a very low prevalence in humans. The parasite could not be detected in the collected lymnaeid snails or on water plants. We also observed distinct differences in the results of copro-microscopy and Ab-ELISA for diagnosing fascioliasis in livestock.

### Low prevalence of human fascioliasis cases despite a high number of historical cases and high prevalence in livestock

Despite the historical data reported by the Center for Disease Control of Nghe An province in Dong Thanh (between 2012 and 2020, 39 cases were reported by the Center for Disease Control of Nghe An province) and the high prevalence in livestock (cattle: 53.74%; buffaloes: 50.45%), we found no human cases based on copro-microscopy and only one case on Ab-ELISA. This discrepancy between the historical prevalence data and our findings may have several reasons. Firstly, changes in farming practices may have significantly contributed to the reduction in *Fasciola* spp. transmission among humans. During our fieldwork, we observed that, while the consumption of raw vegetables remains common, most of these vegetables are cultivated in residents’ private gardens, breaking the life cycle of fascioliasis. These gardens include water plants such as morning glory, rice paddy herb, and fish mint. Another factor that may influence the reporting and detection of *Fasciola* spp. infections is the variability in the diagnostic kits used. In this study, we re-examined 100 of the 1,390 human serum samples using several commercial diagnostic kits for *Fasciola* antibody detection, including the Cortez kit (Human Fasciola IgG ELISA Test Kit, Diagnostic Automation, Cortez Diagnostics Inc., California, USA ), the Khoa Thuong commercial kit from Vietnam (AccuElis Fasciola spp. Detection Kit, KT Biotech, Ho Chi Minh City Vietnam), and the Bio-X kit. Furthermore, we submitted these 100 serum samples to the CDC of Nghe An, which currently uses the Cortez kit as the diagnostic for detecting *Fasciola* spp. antibodies in human sera. Our findings revealed that the Khoa Thuong and Bio-X kits each detected one positive case, aligning with our initial test results. Conversely, when tested at the National Institute of Malariology, Parasitology, and Entomology (NIMPE), the Cortez kit identified two positive cases. Unexpectedly, the CDC Nghe An results indicated seven positive samples. When summarizing the positive cases across the different Ab-ELISA kits, a total of eight positive samples were identified. These eight samples were subsequently subjected to *Fasciola* ES Western Blot IgG kit (LDBIO Diagnostic, 24 AV. Joannes Masset, Lyon, France) which revealed that only one sample consistently tested positive across all methods, corroborating our initial results.

Our findings also indicate that the prevalence of *Fasciola* infection in animals is not a reliable predictor of the prevalence in humans in endemic regions, aligning with previous studies published by Mas-Coma [22] and Tolan [23]. Similarly, a study in Kenya reported no *Fasciola* eggs in human fecal samples, whereas the infection rates in cattle, sheep, and goats were 46.0%, 29.9%, and 24.1%, respectively [24].

### A control program is recommended to reduce the infection rate in cattle and buffaloes

Vietnam currently lacks a coordinated and extensive treatment program for *Fasciola* spp. in livestock. In general, animals are only treated for trematode infections at the owner’s request or when specific symptoms such as weakness, weight loss, or diarrhea are detected. Although certain studies have implemented fascioliasis treatments, these initiatives have typically targeted specific subsets within buffalo and cattle populations rather than including the entire population [25, 26]. In livestock, there is a high risk of infection because the animals are allowed to graze most of the time during the year [7]. With the practice of free-range animal husbandry and the high prevalence of *Fasciola* spp. infection in animals, there is a clear need for a comprehensive fluke control program in the region.

### Age is an important risk-factor of fascioliasis in livestock

Of all the risk-factors that we explored for livestock (host species, age, sex, and source of feed and water), age was the sole risk-factor, with the prevalence of infections increasing over age. Our study demonstrated a positive association between *Fasciola* infection prevalence and intensity and increasing age of livestock. This observation suggests that older animals may have experienced prolonged exposure to the infective stage of the fluke in comparison to their younger counterparts [27]. Older animals, particularly adult buffaloes used in activities such as plowing paddy fields, are more frequently exposed to environments where the intermediate stages of *Fasciola* thrive, such as wet and marshy areas [28]. This frequent exposure accumulates over time, contributing to the increased risk observed in older age groups. This is supported by previous research in Malaysia showing a higher prevalence of infection in cattle older than five years compared to younger animals [29]. Previous research has indicated that the prevalence of *Fasciola* infection is notably higher in older age groups compared to younger ones in buffaloes [30, 31], and in cattle [32, 33].

### Serodiagnosis needs to be carefully deployed across animal hosts

For livestock, we deployed two diagnostic tests with different biomarkers, one that detects eggs in stool (copromicroscopy: Fluke Catcher, Provinos) and one that detects anti-*Fasciola* antibodies in serum (Ab-ELISA: BioX K211, BioX, Rochefort, Belgium). Given the intermittent shedding of eggs by adult flukes in the liver [34], the presence of low numbers of parasites [35], and the long prepatent period (10-14 weeks) [36], it is expected that seroprevalence would be higher than that based on copro-microscopy [37, 38]. Although this expected discrepancy was observed in cattle (copro-prevalence: 50.45% *vs.* Ab-ELISA: 75.90%), in buffaloes the copro-prevalence in buffaloes (53.74%) was significantly higher than the seroprevalence (8.67%). This unexpected finding might be explained by the fact that the Ab-ELISA kit developed by Bio-X for detecting *Fasciola* spp. antibodies in the serum of cattle and sheep might not be suitable for detecting antibodies of *Fasciola* in water buffaloes (*B. bubalis*). Therefore, we recommend that future studies consider the suitability of the Ab-ELISA kit for detecting *Fasciola* antibodies in the serum of water buffaloes.

### Absence of *Fasciola* in snails and water plants

In total, we examined 330 lymnaeid snails across 38 different habitats and 233 water plant samples across 233 households. Despite these sampling efforts, we did not observe any of the samples infected/contaminated with *Fasciola*. Given the high prevalence of fascioliasis in livestock, the reasons for the absence of *Fasciola* might have been due to methodological flaws of the study, including the small sample size, and a less optimal sampling strategy.

Our research primarily focused on understanding the prevalence of *Fasciola* spp. infection in humans rather than in animals. Consequently, we prioritized the collection of aquatic plants commonly consumed raw by households. This focus inherently limited our ability to sample other aquatic plants in the study area that might be contaminated with *Fasciola*, such as those used for livestock feed, including various types of grasses and rice plants. In their 2012 study, Pham et al. detected metacercariae of *Fasciola* spp. on submerged grass in proximity to cattle-grazing areas. Out of the 1,350 samples examined, 255 tested positive for the presence of metacercariae [7].

The timing and scope of snail sampling presented another notable limitation in our study. Snail samples were collected at a single time in May, coinciding with the onset of the rainy season in the region. This period may not be optimal as it likely did not capture the peak snail population, resulting in a total of only 327 lymnaeid snail samples. Previous studies in Vietnam have reported *Fasciola* infection rates in snails ranging from 0% to 1.8% [39]. Therefore, the relatively low number of samples collected in our study limits the likelihood of detecting positive cases. Similar results were observed in a comparable study conducted in the Philippines, which found no positive cases of *Fasciola* spp. among 750 lymnaeid snail samples [40]. Our sample size was significantly smaller than those reported in other studies, such as the study by Quy et al., which examined 2,269 snails and identified a *Fasciola* infection rate of 0.5% [14]; the study by Bui et al, which analyzed 24,488 snails with an infection rate ranging from 0% to 0.7% [41]; and the study by Suhardono et al, which investigated 10,159 snails and found an infection rate of 1.8% [42]. To enhance the detection of *Fasciola* spp. and to gain a comprehensive understanding of its transmission in snails, it is common for studies to collect snail samples at different times throughout the year [33, 40, 43]. These longitudinal studies provide more representative data on the prevalence and transmission dynamics of *Fasciola* spp. in snails over time.

## Conclusion

We confirmed a high prevalence of fascioliasis in livestock in a North Vietnamese rural area, with older animals being at higher risk of infection and in need of treatment. Our findings in humans were not in line with historical data and underscore potential behavioral changes that impede (zoonotic) disease transmission and the need for more reliable diagnostic tests in national and provincial laboratories to inform health decision-making. We also highlight the need for an Ab-ELISA that is validated for use in buffaloes, and that further research is required to improve both sampling and diagnostic means to demonstrate infections in snails and water plants

## Data Availability

Data will be made available on acceptance of the manuscript.

## 5. Acknowledgments

This study was supported by the EmFaVie project, funded by the Flemish Interuniversity Council - University Development Cooperation (VLIR-UOS, grant number VN2020SIN317A103, granted to BL and DDT), and the FasciCoM project, jointly funded by the Research Foundation - Flanders (FWO, grant number G0E2921N, granted to BL) and the Vietnamese National Foundation for Science and Technology Development (NAFOSTED, grant number FWO.108.2020.01, granted to DDT). The funders had no role in study design, data collection and analysis, decision to publish, or preparation of the manuscript. We thank the European Virus Archive GLOBAL (EVA-GLOBAL) project for providing PhHV-1, which received funding from the European Union’s Horizon 2020 research and innovation program under grant agreement No 871029. We would like to extend our special thanks to all the study participants involved in this survey. We are particularly grateful to our field team members, Nguyen Duc Thuy, Tran Thi Tuyen, Nguyen Huu Truong, and Dinh Cong Thinh, for their dedicated support during the fieldwork. We also appreciate the assistance provided by the Nghe An Province Center for Disease Control, the Yen Thanh Health District Center, and the health stations in Dong Thanh commune.

## 7. Supporting information

Info S1. Individual questionnaire for human participants

Info S2. Household questionnaire for head of household

Info S3. Instructions for collecting freshwater plants

Info S4. Standard operating procedure for stool sample examination

Info S5. Standard operating procedure for serum antibody detection (Ab-ELISA)

